# Stimulus-induced Gamma rhythms are weaker in human elderly with Mild Cognitive Impairment and Alzheimer’s Disease

**DOI:** 10.1101/2020.06.24.20139113

**Authors:** Dinavahi V. P. S. Murty, Keerthana Manikandan, Ranjini Garani Ramesh, Simran Purokayastha, Bhargavi Nagendra, M. L. Abhishek, Aditi Balakrishnan, Mahendra Javali, Naren Prahalada Rao, Supratim Ray

## Abstract

Alzheimer’s Disease (AD) in elderly adds substantially to socio-economic burden necessitating early diagnosis. While recent studies in rodent models of AD have suggested diagnostic and therapeutic value for gamma rhythms in brain, the same has not been rigorously tested in humans. We recruited a large population (N=247; 106 females) of elderly (>49 years) individuals from the community, who viewed large gratings that induced strong gamma oscillations in their electroencephalogram (EEG). These individuals were classified as healthy (N=227), mild-cognitively-impaired (MCI; 14) or AD (6) based on clinical history and Clinical Dementia Rating scores. Surprisingly, stimulus-induced gamma rhythms, but not alpha or steady-state-visually-evoked-responses, were significantly lower in both MCI and AD patients compared to their age and gender matched controls. This reduction was not due to differences in eye movements or baseline power. Our results suggest that gamma could be used as potential diagnostic tool for MCI/AD in humans.

**One Sentence Summary:** A large double-blinded EEG study suggests that narrow-band visual gamma rhythms are weaker in MCI/AD patients compared to cognitively healthy controls.

## Introduction

Alzheimer’s Disease (AD) is a predominant cause of dementia (decline in cognitive abilities) of old age and substantially contributes to the socio-economic burden in the geriatric population, necessitating early diagnosis. Advances in our understanding of cellular pathology of AD in rodent models and its link to gamma rhythms in brain has spurred interest to investigate diagnostic and therapeutic potential of gamma rhythms in AD and other forms of dementia *(1, 2)*.

Gamma rhythms are narrow-band oscillations in brain’s electrical activity with center frequency occupying ∼30-80 Hz frequency range *(3)*. These are suggested to be generated from excitatory-inhibitory interactions of pyramidal cell - interneuron networks *(4)* involving parvalbumin (PV) and somatostatin (SOM) interneurons *(5–7)*. These have been proposed to be involved in certain cognitive functions like feature binding *(3)*, attention *(8–10)* and working memory *(11)*.

Some studies have reported abnormalities in gamma linked to interneuron dysfunction in AD. For example, Verret et al. *(12)* reported PV interneuron dysfunction in parietal cortex of AD patients and hAPP mice (models of AD). They found aberrant gamma activity in parietal cortex in such mice. Further, some recent studies have suggested therapeutic benefit of entraining brain oscillations in gamma range in rodent models of AD. For example, Iaccarino *et al. (13)* suggested that visual stimulation using light flickering at 40 Hz entrained neural activity at 40 Hz and correlated with decrease in Aβ amyloid load in visual cortices of 5XFAD, APP/PS1 mice models of AD. Based on such reports in rodents in both visual and auditory modalities, some investigators have suggested a paradigm termed GENUS (gamma-entrainment of neural activity using sensory stimuli) and have claimed to show neuro-protective effects in rodent models of AD *(14, 15)*.

Recent studies in human EEG *(16, 17)* and MEG *(18)* have reported existence of two gamma rhythms (slow: ∼20-34 Hz and fast: ∼36-66 Hz) in visual cortex, elicited by Cartesian gratings. Age-related decline in power and/or frequency of these stimulus-induced gamma rhythms has been shown in cognitively healthy subjects *(17, 19)*. However, abnormalities in such stimulus-induced visual narrow-band gamma rhythms in human patients of MCI (Mild Cognitive Impairment, a preclinical stage of dementia *(20–22)*) or AD have not been demonstrated till date.

We addressed this question in the present double-blind case-control EEG study involving a large cohort of elderly individuals (N=247; 106 females, all aged >49 years) recruited from urban communities in Bangalore, India. These were classified as healthy (N=227; *see 17*), MCI (14) or AD (6) based on clinical history and Clinical Dementia Rating (CDR; *23, 24*) scores. We studied narrow-band gamma rhythms induced by full screen Cartesian gratings in all these participants. We also examined steady-state visually evoked potentials (SSVEPs) at 32 Hz in a subset of these participants (9 MCI, 3 AD and 76 healthy controls). We monitored eyes using infrared eye tracker to rule out differences in gamma power due to potential differences in eye position or microsaccade rate *(25)*.

## Results

First, we examined how the two gamma rhythms differed in cases as compared to their healthy age and gender matched controls using full-screen static sinusoidal grating stimuli presented on a computer monitor. Out of the 227 cognitively healthy participants, we selected 114 controls who were age (± 1 year) and gender matched with their respective cases (MCI/AD) for analysis. One AD patient (A1 in Supplementary Figure 1, 92 years old male) did not have any matching control.

We averaged spectral data for all analyzable bipolar electrodes *(as described in 17)* from 10 occipital and parieto-occipital pairs (marked in black enclosures in Figure 1d; see SI Methods). Figure 1a shows the median stimulus-induced change in PSDs for 20 cases and 114 age and gender matched controls pooled together (light shaded regions show ±SD of median after bootstrapping for 10,000 iterations). While both slow and fast gamma ‘bumps’ were conspicuously visible for case as well as control groups, power in both slow gamma (20-34 Hz) and fast gamma (36-66 Hz) ranges (but not alpha, 8-12 Hz range) was significantly lower in the case group compared to the control group (Kruskal-Wallis Test, significance as shown in Figure 1a). This could also be seen in the median time-frequency change in power spectrograms (baseline: −500-0 ms of stimulus onset) for cases and controls in Figure 1b.

**Fig 1.**
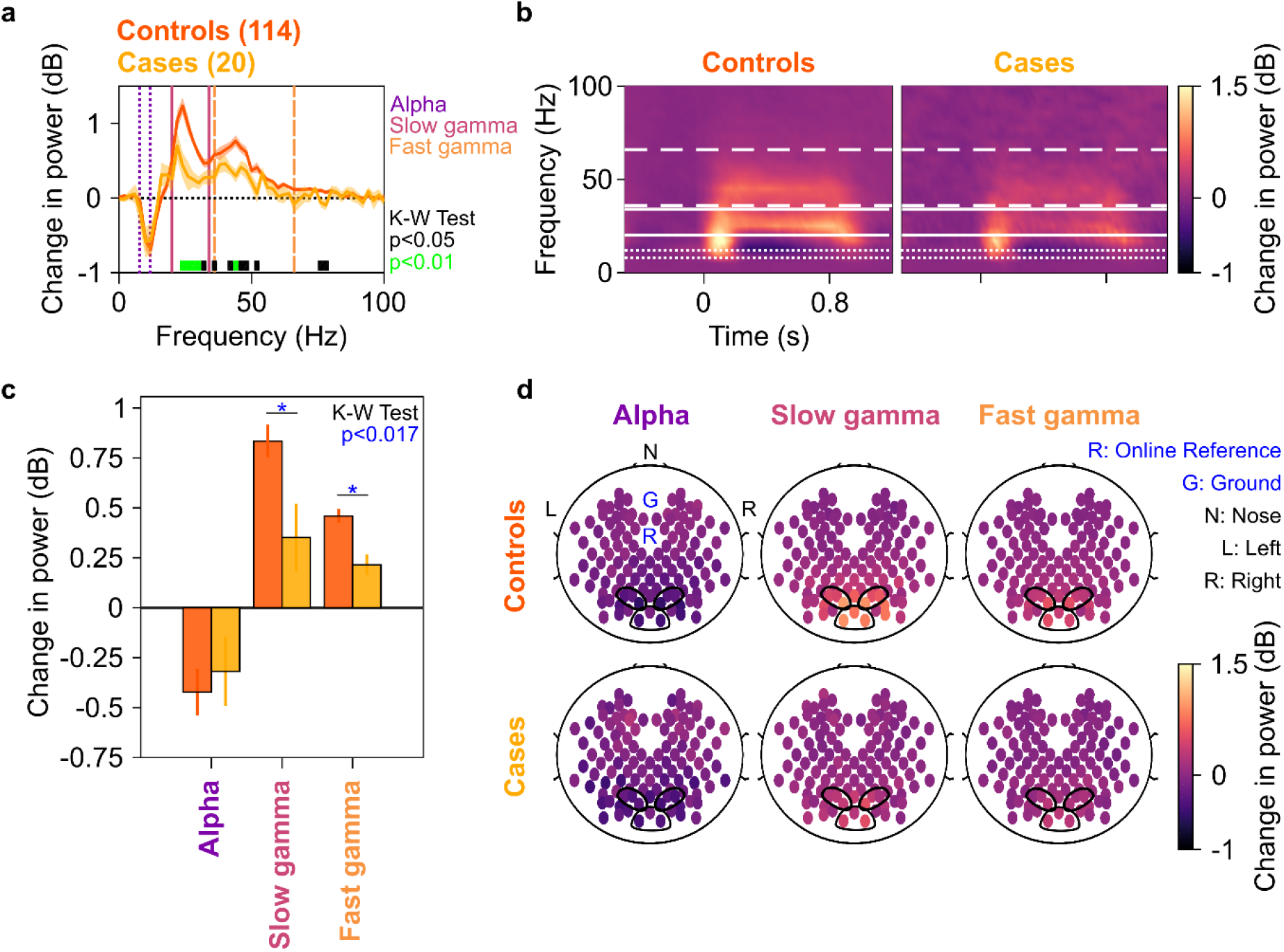
Alpha, slow and fast gamma in cases and controls. a) Change in power spectral densities (PSD) for 20 cases (yellow) and 114 controls (dark orange). Solid PSD traces indicate median, and shaded regions indicate ±SD from medians after bootstrapping over 10,000 iterations. Vertical lines represent alpha (8-12 Hz, violet), slow (20-34 Hz, pink) and fast gamma (36-66 Hz, orange). Colored bars at the bottom represent significance of differences in medians (Kruskal-Wallis test, black: p<0.05 and green: p<0.01; not corrected for multiple comparisons). b) Median change in power spectrograms for cases (right) and controls (left). White horizontal lines represent alpha (dotted), slow gamma (solid) and fast gamma (dashed) bands. c) Median change in power in alpha, slow gamma and fast gamma bands for cases (yellow) and controls (dark orange). Error bars indicate ±SD from medians after bootstrapping over 10,000 iterations. Blue asterisks represent significance of differences in medians (K-W test, p<0.017 after Bonferroni-adjustment) d) Average scalp maps of 112 bipolar electrodes (disks) for cases (bottom row) and controls (top row) for alpha (left), slow gamma (middle) and fast gamma (right). Color of disks represents change in power in respective frequency bands. Electrode groups used for calculation of band-limited power are enclosed in black.

Change in band-limited power was lesser for both gamma bands in the case group compared to the control group (Figure 1c, KW test, χ^2^(133)=6.14, p=0.013 for slow and χ^2^(133)=8.00, p=0.005 for fast gamma, both significant at a Bonferroni-adjusted significance level of 0.05/3=0.017). However, this was not true for alpha range (Figure 1c, KW test, χ^2^(133)=0.11, p=0.741). Figure 1d shows the median scalp maps *(EEGLAB, 26*, see SI Methods) of change in band-limited power across 112 bipolar electrode pairs (shown as discs) for alpha, slow and fast gamma bands. We observed that stimulus-induced change in power across all 3 bands was most prominent in the 10 electrode pairs as above. However, this change in power was less in the case group compared to the control group in slow and fast gamma bands (but not alpha band) as noted in Figure 1c. These trends remained similar if we compared the MCI and AD groups separately with their respective controls (Supplementary Figure 1). Further, these trends did not differ qualitatively if we removed participant A1 (who did not have a corresponding control) from analysis.

To confirm these group-level results at individual level, we compared the change in band-limited power for each case with the median change in power of their corresponding age and gender matched controls. Figure 2 shows scatter plots of band-limited power in alpha, slow and fast gamma for 19 cases (excluding A1), and the median change in power for their corresponding controls. Only 3 MCI patients (and 0 AD patients) had higher slow gamma than their controls. Similarly, only 2 MCI and 1 AD patients had higher fast gamma than their controls. However, for alpha, the scatter was symmetrical across the identity line. This shows that most of the controls had higher slow/fast gamma (and not alpha) than the cases, thus corroborating the results observed in Figure 1.

**Fig 2.**
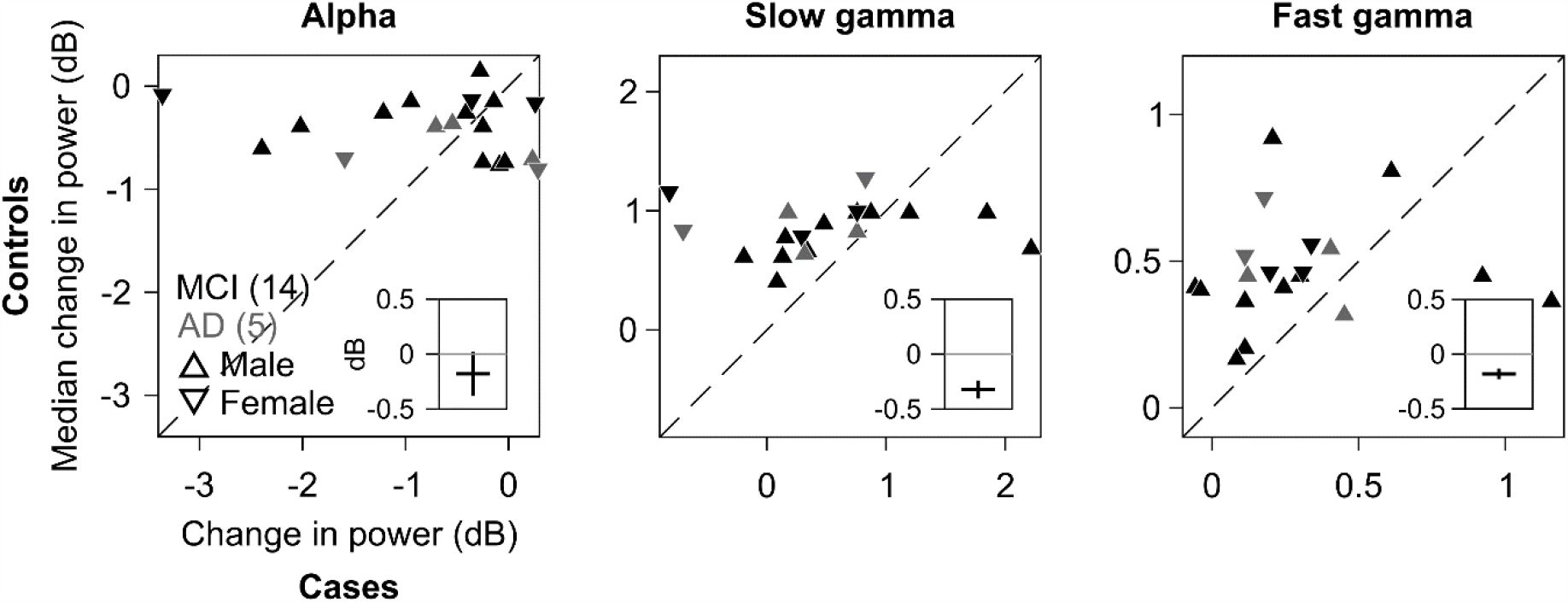
Change in alpha, slow and fast gamma power for each case compared to corresponding controls. Scatter plots showing change in power for each of the 19 cases (abscissa) and median change in power for corresponding controls (ordinate) in alpha (right), slow gamma (middle) and fast gamma (left). MCI are plotted in black and AD in gray. Triangles pointing upwards indicate males and those downwards indicate females. 1 case (92 years, male, AD) did not have any age and gender matched control and was discarded. Dashed line represents identity line. Insets: differences in change in power values for 19 cases and their corresponding controls, plotted as median ± SD (error bar, boot strapped over 10,000 iterations).

To statistically test this, we calculated the difference between change in power for each case and the median change in power of their corresponding controls, separately for alpha, slow gamma and fast gamma bands. This yielded a set of 19 differences for each of the three bands. The median difference was significantly less than zero for slow and fast gamma (left-tailed paired Wilcoxon signed-rank test, Z=-2.35/-2.27, p=0.009/0.01 for slow/fast gamma respectively), but not alpha (Z=-0.95, p=0.17). Results remained similar when we randomly chose only 1 control per case (median p-values over 10,000 iterations: 0.162/0.031/0.048 for alpha/slow/fast gamma). In addition, median difference ± bootstrapped SD of median difference, over 10,000 iterations, did not include 0 dB for either gamma bands but did so for alpha (as depicted in the insets in Figure 2). These trends did not differ qualitatively when we reanalysed the data after removing 4 MCI and 1 AD patients who had negative change in either slow or fast gamma power (Supplementary Figure 2a). Further, to minimise error during clinical diagnosis, we called for a review of the diagnosis of MCI/AD patients by an expert panel that operationalised NIA-AA criteria to study setting (see SI Methods). This panel revised the diagnosis of 2 MCI patients (M4 and M8) as healthy. Trends did not differ when we removed these 2 patients from analysis (Supplementary Figure 2b). These results indicated that both slow and fast gamma were weaker in the cases compared to the controls.

We found that the cases and their respective controls had comparable eye-movement and microsaccade profiles, and similar pupillary reactivity to stimulus presentation (measured as coefficient of variation of pupil diameter across time, *see 17*). Further, the trends described in Figures 1 and 2 did not change when we reanalysed the data after removing stimulus repeats containing microsaccades (*17, 27*, see SI Methods) from analysis (Supplementary Figure 3). Finally, the cases and controls had comparable PSDs and slopes of PSDs in the baseline condition (Supplementary Figure 4). We thus ruled out the biases introduced by these non-neural variables in our analyses.

We next tested whether power of steady-state visually evoked potentials (SSVEPs) in gamma range also decreased in the case group as compared to the control group. We tested for SSVEPs at 32 Hz by presenting full-screen gratings that phased-reversed at 16 Hz. 17 of the 20 cases participated in this study, out of which data of only 12 could be analysed (9 MCI, 3 AD; data from 5 cases were discarded due to noise, see SI Methods for details). These 12 cases had a total of 76 healthy age and gender matched controls. Figures 3a and 3b show change in power spectral density plots (in 250-750 ms window of stimulus onset) and change in power time-frequency spectrograms (from a baseline period of −500-0 ms of stimulus onset, same as in Figures 1a and 1b) for these 12 cases and 76 healthy age and gender matched controls, pooled together. Figures 3c and 3d show bar plots and scalp maps for 112 bipolar electrodes for change in SSVEP power at 32 Hz (during 250-750 ms of stimulus onset from a baseline of −500-0 ms, same as in Figures 1c and 1d) for control and case groups, respectively. We observed a modest reduction in SSVEP power at 32 Hz in the case group as compared to the control group (Figures 3a and 3c), but this decreasing trend was not significant (K-W Test, significance at each frequency of the change in power spectra is shown in Figure 3a. Significance at 32 Hz: χ^2^(87)=0.50, p=0.48). This contrasted with trends for gamma power: we reanalysed data for slow and fast gamma power (as in Figure 1) with the same set of 12 cases and 76 controls (Supplementary Figures 5a and 5b, same format as Figures 1a and 1c respectively). We found that cases had significantly less fast gamma power compared to controls (KW test, χ^2^(87)=6.03, p=0.014, significant at a Bonferroni-adjusted significance level of 0.05/3=0.017). Slow gamma also showed a decreasing trend like in Figure 1, albeit the difference was not significant (χ^2^(87)=0.85, p=0.35). Alpha followed a similar insignificant trend as in Figure 1 (χ^2^(87)=0.55, p=0.45).

**Fig 3.**
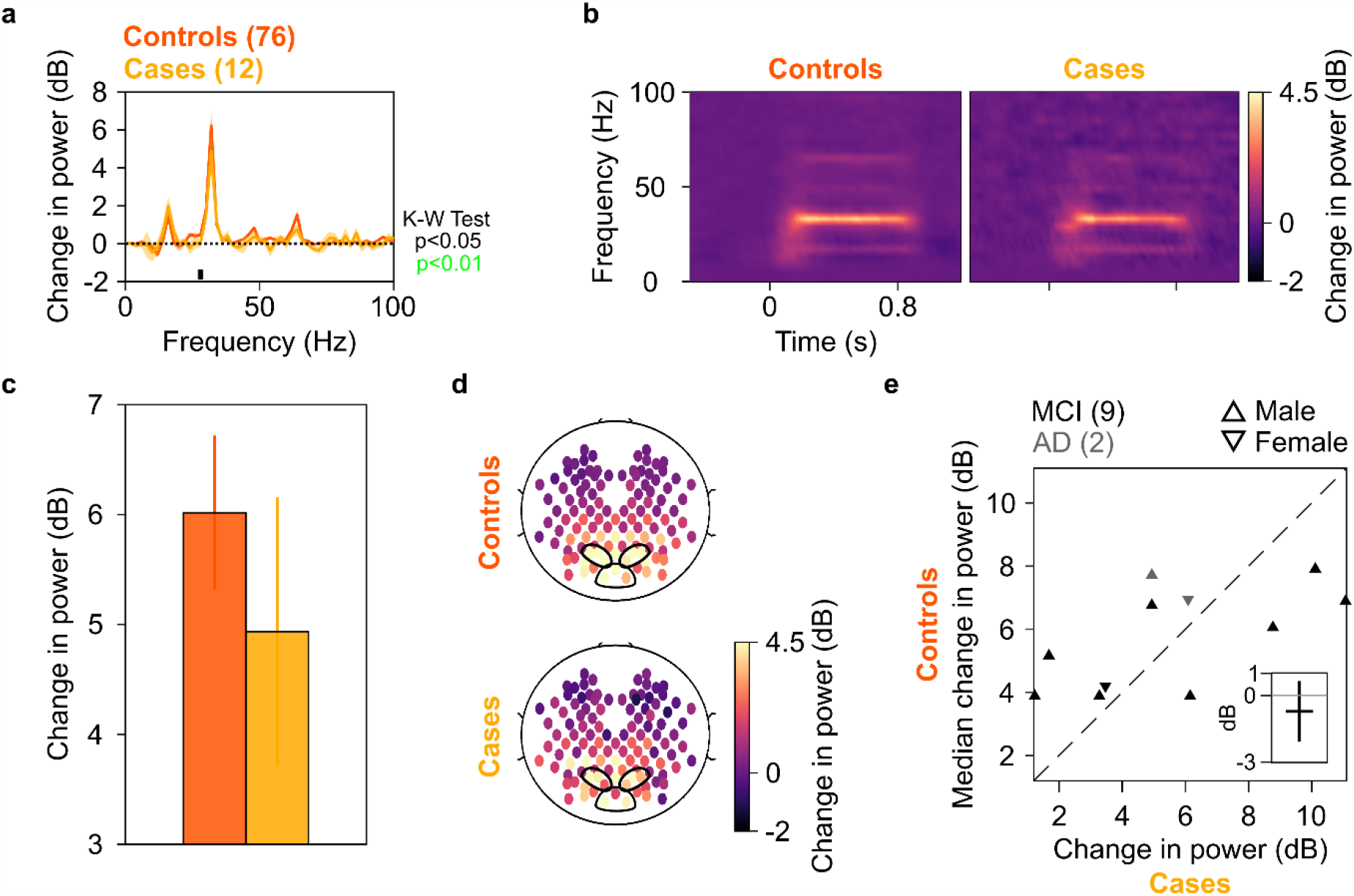
SSVEP at 32 Hz in cases and controls. Median change in PSD (solid traces in (a)) and median change in power spectrograms in (b) for 12 cases (N = 9/3 for MCI/AD) and 76 controls. Shaded regions in (a) indicate ±SD from medians after bootstrapping over 10,000 iterations. c) Median change in SSVEP power at 32 Hz for cases (yellow) and controls (dark orange). Error bars indicate ±SD from medians after bootstrapping over 10,000 iterations. d) Average scalp maps of 112 bipolar electrodes (disks) for cases (bottom row) and controls (top row) for change in power at 32 Hz. Same format as in Figure 1d. e) Scatter plots showing change in SSVEP power at 32 Hz for each of the 11 cases (abscissa; N = 9/2 for MCI/AD) and median change in power for corresponding controls (ordinate). 1 case (A1, 92 years, male, AD) did not have any age and gender matched control and has been discarded. Same format as in Figure 2.

We also repeated the analysis at individual level as in Figure 2. Figure 3e shows scatter plot for change in SSVEP power at 32 Hz for each of the 11 cases (participant A1 discarded) and the median change in SSVEP power for the corresponding controls, same format as in Figure 3. We observed that there was a lot of scatter around the identity line, suggesting that SSVEP power was not less in cases compared to their controls, in contrast to slow and fast gamma power. To quantify the significance of our observation, we calculated differences of change in SSVEP power at 32 Hz for 11 cases and median change in SSVEP power of corresponding controls. The median difference was not significantly less than 0 dB (left-tailed paired Wilcoxon signed-rank test, Z=-0.22, p=0.4). Moreover, the median difference ±SD (after bootstrapping over 10,000 iterations) included 0 dB mark (inset in Figure 3e), suggesting that these differences were not significantly different from 0 dB. The trends for alpha/slow/fast gamma for these 11 cases and their respective controls were comparable to that discussed in Figure 2 (Supplementary Figure 5c, same format as Figure 2). To conclude, change in SSVEP power at 32 Hz for cases was comparable to that of their controls, like alpha but unlike slow and fast gamma oscillations.

## Discussion

Stimulus-induced change in power of both narrow-band slow and fast gamma oscillations reduced in elderly patients with clinical MCI/AD compared to their age and gender matched healthy controls. We removed or ruled out possible biases due to peripheral ocular factors or overall baseline noise of the PSDs to further strengthen the results. In contrast to gamma, we did not find any significant reduction in stimulus-induced alpha suppression or SSVEP at 32 Hz in cases.

Previous studies have suggested abnormalities in spontaneous and evoked activity in gamma band in various disorders. Some studies have also explored abnormalities in other oscillatory bands such as alpha as well as functional connectivity across brain regions in such disorders. Examples include autism *(28, 29)*, schizophrenia *(30, 31)* and AD and other dementias *(32–34)*, etc. However, this is the first study in our knowledge that found abnormalities in visual narrow-band gamma oscillations elicited by Cartesian gratings in human EEG in MCI/AD.

Our sample is a representative of urban population in India as we adopted community-based sampling instead of hospital-based sampling. Importantly, out of the 257 participants that we collected data from (247 used for analysis plus 10 participants whose data was noisy and thus rejected, see SI Methods), there were 15 MCIs (5.8%) and 6 AD patients (2.3%). These figures match closely to the previously reported prevalence of MCI and AD in India *(22, 35, 36)*. The main strength of our study is that within this sample, most cases (70%) had MCI, a condition that is conceptualized as intermediate stage between normal aging and AD. Criteria used to diagnose MCI are not strong and hence there is a need for a valid biomarker. Our study highlights the potential use of gamma oscillations in EEG in that direction.

Moreover, we had limited our analyses to sensor (electrode) level instead of reconstructing the neural sources and performing analyses at that level. This has allowed us to present a diagnostic technique that is easy to replicate in a clinical setting. Furthermore, as our metrics were derived directly from neural activity, these could serve as a more objective assessment of the clinical status of the individual. Future studies should examine if different other stimulus paradigms like annular gratings *(37)* and drifting gratings *(38)* could add to the robust evidence that is presented in this study.

Gamma rhythms are generated by excitatory-inhibitory interactions in the brain *(4)*. These interactions could be influenced by many structural factors *(39)* that could get abnormal in AD (such as cortical thinning and atrophy; *see 40, 41*). However, how such structural derailments influence gamma recorded over scalp is unknown. A few studies in MEG had reported significant positive correlations between gamma frequency and cortical thickness as well as volume of cuneus *(19)* and thickness of pericalcarine area *(42)*. However, such results could not be replicated in later studies *(43)* and have been shown to be confounded by age *(44)*. Age is as a common factor that influences both macroscopic structure *(45–47)* as well as gamma power and frequency *(17)*.

Our main aim in this study was to examine the potential of gamma as a biomarker. However, as structural changes in AD brain are more evident and drastic compared to cognitively healthy aging *(40, 41)*, future studies should examine correlations of gamma power/frequency and macroscopic structure such as cortical thickness in healthy/MCI/AD individuals while controlling for age. Moreover, as gamma rhythms have been correlated with many higher level cognitive functions such as attention, working memory, etc. (see Introduction), attempts must be made to extend and validate these findings in clinical populations, such as AD.

Some investigators have suggested neuroprotective effects of entraining neural oscillations using flickering light/sound in gamma frequency range (analogous to our SSVEP paradigm), in rodent models of AD *(14, 15, 48)*. However, we did not find any significant trend for SSVEP at 32 Hz in cases compared to controls, unlike our observations with narrow-band gamma. There could be several reasons for these differences. First, it is possible that entrainment of neural oscillations to visual stimulation in gamma frequency range gets deranged only in advanced stages of AD (as in the rodent models of *13*). It may thus have therapeutic benefit but may not reflect as abnormal on testing early on (as in our case). Second, as discussed in the SI Methods, SSVEP study was always done at the end of the experiment in our study and the total number of stimulus repeats were much less than the gamma study.

To conclude, stimulus-induced change in visual narrow-band gamma power has the potential to be a simple, low-cost, easy to replicate and objective biomarker for diagnosis of MCI/AD. How this gamma-based biomarker compares against other methods used in diagnosis (MRI, PET, cognitive tests etc.), whether addition of this biomarker to other standard methods improves the overall diagnosis, and the specificity of this biomarker for AD compared to other causes of dementia remain open questions that will require further research.

## Materials and Methods

### Participants

We recruited 257 elderly participants (109 females) aged 50-92 years from the Tata Longitudinal Study of Aging (TLSA) cohort from urban communities in Bangalore between July 2016 and July 2019. They were clinically diagnosed by psychiatrists (authors BN/AML) and/or a neurologist (author MJ) as cognitively healthy (N=236), MCI (15) or AD (6) through clinical history and a semi-structured clinical interview (Clinical Dementia Rating or CDR; see Table 1). 5 out of the 6 AD patients were directly referred to the study by the neurologist. Diagnosis of all MCI/AD patients was further reviewed by a panel of 4 experts for consensus (see SI Methods). We discarded data of 10 participants due to noise (see Artifact Rejection section in SI Methods). We were thus left with 227 healthy participants, 14 MCI and 6 AD patients for analysis. For the purpose of this study, we called the MCI/AD patients as cases (N=20) and their respective age and gender matched healthy participants as controls.

**Table 1.**
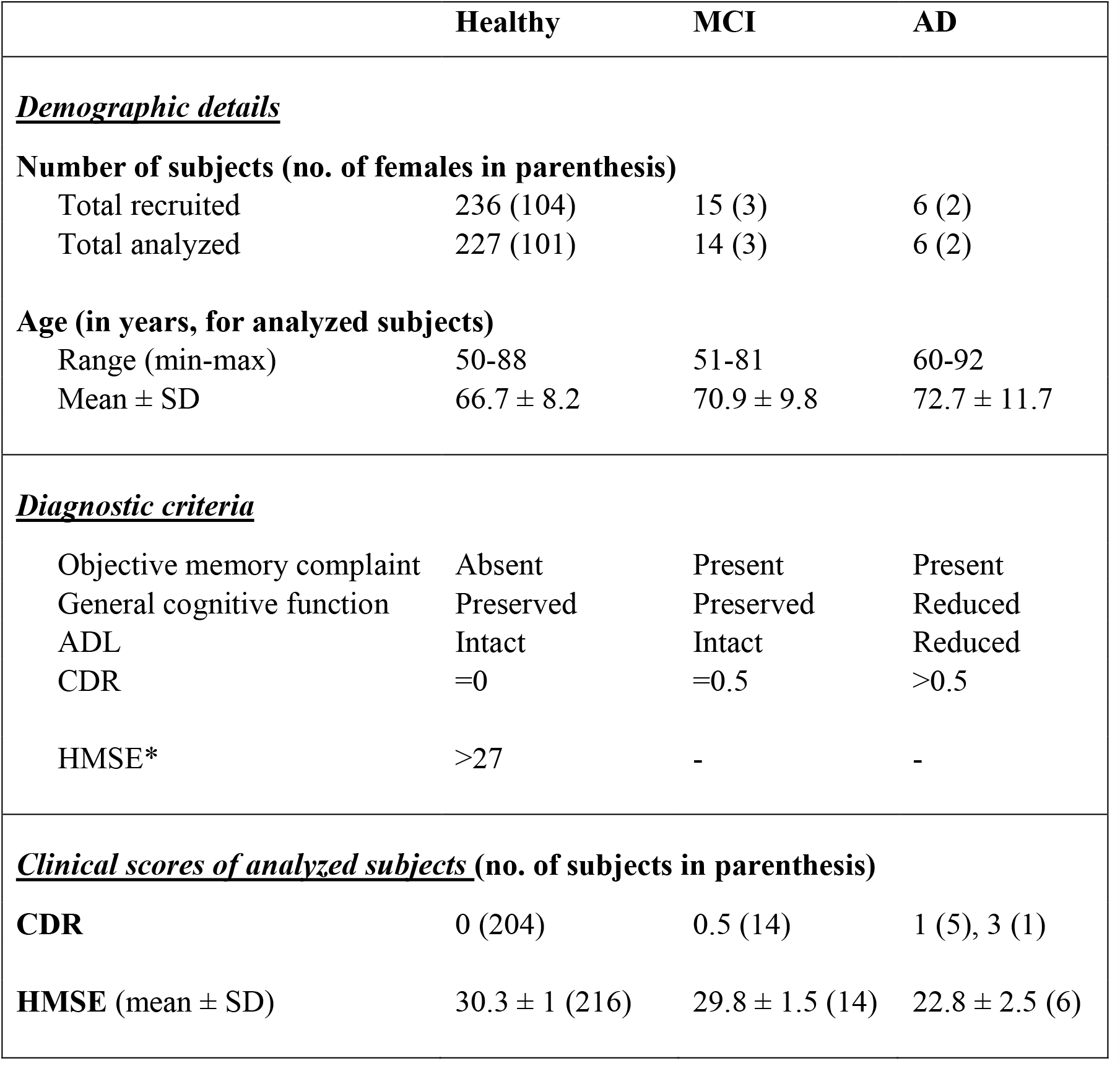
Demographic and clinical details for participants. MCI: Mild Cognitive Impairment; AD: Alzheimer’s disease; ADL: Activities of Daily Living; CDR: Clinical Dementia Rating *(23, 24)*; HMSE: Hindi Mental State Examination *(51)*. *HMSE was used as a diagnostic criterion only if CDR score was unavailable and clinical testing did not indicate any sign of dementia.

|  | Healthy | MCI | AD |
| --- | --- | --- | --- |
| **<u>Demographic details</u>** |  |  |  |
| **Number of subjects (no. of females in parenthesis)** |  |  |  |
| Total recruited | 236 (104) | 15 (3) | 6 (2) |
| Total analyzed | 227 (101) | 14 (3) | 6 (2) |
| **Age (in years, for analyzed subjects)** |  |  |  |
| Range (min-max) | 50-88 | 51-81 | 60-92 |
| Mean ± SD | 66.7 ± 8.2 | 70.9 ± 9.8 | 72.7 ± 11.7 |
| **<u>Diagnostic criteria</u>** |  |  |  |
| Objective memory complaint | Absent | Present | Present |
| General cognitive function | Preserved | Preserved | Reduced |
| ADL | Intact | Intact | Reduced |
| CDR | =0 | =0.5 | >0.5 |
| HMSE* | >27 | - | - |
| **<u>Clinical scores of analyzed subjects</u> (no. of subjects in parenthesis)** |  |  |  |
| **CDR** | 0 (204) | 0.5 (14) | 1 (5), 3 (1) |
| **HMSE** (mean ± SD) | 30.3 ± 1 (216) | 29.8 ± 1.5 (14) | 22.8 ± 2.5 (6) |

All participants reported normal or corrected-to-normal vision and were instructed to wear spectacles if prescribed earlier. They participated in the study voluntarily and were monetarily compensated for their time and effort. We obtained informed consent from all participants before the experiment. The Institute Human Ethics Committees of Indian Institute of Science, NIMHANS, and M S Ramaiah Hospital, Bangalore approved all procedures. This article is in compliance with the STROBE statement *(49)*.

### Experimental setting, task and analysis

Experimental setting, task, artefact rejection and data analysis were as described in Murty et al. *(17)*. Briefly, participants performed a visual fixation task in which we presented a series of full screen static sinusoidal luminance gratings on a computer monitor. The gratings were presented for 800 ms (with interstimulus interval of 700 ms) at one of three spatial frequencies (SFs): 1, 2, and 4 cycles per degree (cpd) and four orientations: 0°, 45°, 90° and 135° for the Gamma experiment, chosen pseudorandomly. We also presented gratings (with one SF and orientation combination that showed high change in gamma power for each participant during preliminary analysis performed for Gamma experiment) counter-phasing at 16 cycles per second (cps), to test for SSVEPs at 32 Hz in the SSVEP experiment. Both the experiments were performed in a single recording session lasting ∼30 minutes. We recorded 64-channel EEG using active electrodes (international 10-10 system of electrode placement) and BrainAmp DC (Brain Products GmbH). We also recorded eye data using an infrared eye-tracker EyeLink 1000 (SR Research Ltd) for all participants (except for one each in healthy/MCI/AD categories). After rejecting data from fixation breaks and noisy stimulus repeats/electrodes/participants from analysis, we pooled data across the following bipolar occipital and parieto-occipital pairs (that had analyzable data): PO3-P1, PO3-P3, POz-PO3, PO4-P2, PO4-P4, POz-PO4, Oz-POz, Oz-O1, Oz-O2. We used multitaper spectral analysis method (using one taper) to estimate PSD and time-frequency spectrograms, using Chronux toolbox (*50*, http://chronux.org/, RRID:SCR_005547). We chose −0.5-0 s as baseline period and 0.25-0.75 s as stimulus period; PSDs therefore had a frequency resolution of 2 Hz. We chose a moving window of size 250 ms and step size of 25 ms for the spectrograms, giving a frequency resolution of 4 Hz. We refer to supplementary (SI) methods for more details.

### Statistical analysis

All our statistical interpretations were based on non-parametric tests on medians using either Kruskal-Wallis Test or Wilcoxon signed-rank test (for paired comparisons). We used Bonferroni correction for multiple tests/comparisons wherever necessary.

## Author contributions: DVPSM

Conceptualization; Data curation; Formal analysis; Investigation; Methodology; Visualization; Roles/Writing - original draft; Writing - review & editing; **KM:** Formal analysis; Data curation; Investigation; **RGR, SP, BN, AML, AB:** Data curation; Investigation; **MJ:** Validation; Writing - review & editing; **NPR:** Conceptualization; Data curation, Funding acquisition; Methodology, Project administration; Resources, Supervision; Validation; Writing - review & editing; **SR:** Conceptualization; Data curation; Formal analysis; Funding acquisition; Methodology; Project administration; Resources; Supervision; Validation; Visualization; Roles/Writing - original draft; Writing - review & editing

## Data and materials availability

The EEG data presented here is recorded as part of a large multi-investigator project that involved several other experiments and measurements like psychophysics, fMRI, PET, etc, some of which are still in progress. Hence, the data would be made available to readers upon request and publicly available at a later time, according to the policies of the project. All spectral analyses were performed using Chronux toolbox (version 2.10), available at http://chronux.org.

## Supplementary Information

Title: Stimulus-induced Gamma rhythms are weaker in human elderly with Mild Cognitive Impairment and Alzheimer’s Disease

Authors: Dinavahi V. P. S. Murty, Keerthana Manikandan, Ranjini Garani Ramesh, Simran Purokayastha, Bhargavi Nagendra, Abhishek M. L., Aditi Balakrishnan, Mahendra Javali, Naren Prahalada Rao and Supratim Ray.

## SI Methods

### Consensus diagnosis

As in Table 1, our major criteria to diagnose MCI/AD were clinical history and CDR which was administered by a single clinician (neurologist or psychiatrist). However, to minimize observer bias, we followed the recommendations made in SAGES study *(52)* to arrive at a consensus diagnosis for each MCI/AD patient. We constituted a panel with four members, consisting of one neurologist (author MJ), two psychiatrists (authors AML and NPR) and one psychologist (author AB, an additional expert who did not participate in the single clinician diagnosis before). These members independently reviewed data variables available in TLSA (Tata Longitudinal Study of Aging) cohort to operationalize the NIA-AA criteria *(53)* for probable AD and MCI. For each of the NIA-AA criteria, they operationalized an equivalent TLSA criterion (Supplementary Tables 1-3). Wherever longitudinal data was available, they considered trends over time for diagnosis.

They labelled a subject as MCI/AD only if all 4 of them concurred with the diagnosis. When they could not achieve at a consensus for any subject in the first instance (6 subjects), they used Delphi method *(54, 55)* till all of them agreed upon the diagnosis. Briefly, the members were informed of the discrepancy within the panel, who then discussed among themselves and rated again. This process was iterated till all 4 members came to a consensus. Although they used this stringent approach to confirm the previous diagnoses, they reclassified as healthy only 2 subjects previously classified as MCI (M4 and M8, see Supplementary Figure 1). They confirmed and retained initial diagnosis for rest of the 12/14 MCI and 6/6 AD patients. The analyses results for these patients are presented in Supplementary Figure 2b.

### Experimental setup and task

Experimental setup, EEG recordings and analysis were same as what we had described in our previous study *(17)*. Briefly, we recorded raw EEG signals from 64 active electrodes using BrainAmp DC (Brain Products GmbH) according to the international 10-10 system, referenced online at FCz. We filtered raw signals online between 0.016 Hz and 1000 Hz and sampled at 2500 Hz. We rejected electrodes whose impedance was more than 25 KΩ (4.0% and 2.5% for healthy subjects and cases respectively). Impedance of the final set of electrodes was 5.5±4.2 and 5.2±4.3 KΩ for healthy subjects and cases respectively.

All subjects sat in a dark room in front of an LCD screen with their head supported by a chin rest. The screen (BenQ XL2411, resolution: 1280 × 720 pixels, refresh rate: 100 Hz) was gamma-corrected and placed at a mean±SD distance of 58.1±0.8 cm from the subjects (53.8-61.0 cm) according to their convenience (thus subtending a width of at least 52° and height of at least 30° of visual field for full-screen gratings). We calibrated the stimuli to the viewing distance in all cases.

Subjects performed a visual fixation task. Every trial started with the onset of a fixation spot (0.1°) at the center of the screen on which the subjects had to maintain fixation. After an initial blank period of 1000 ms, a series of stimuli (2 to 3 sinusoidal luminance gratings presented full screen at full contrast) were randomly shown for 800 ms each with an inter-stimulus interval of 700 ms. For the main “Gamma” experiment, these were presented at three spatial frequencies (SFs): 1, 2, and 4 cycles per degree (cpd) and four orientations: 0°, 45°, 90° and 135°. Subjects performed this task in 2-3 blocks (total 543 blocks across 257 subjects) during a single session according to their comfort.

We also tested 32-Hz SSVEPs on these subjects in the SSVEP experiment. Gratings (with one SF and orientation combination that showed high change in slow and fast gamma power for each subject during preliminary analysis performed during the session) counter-phased at 16 cycles per second (cps) in a similar stimulus presentation paradigm as described above, randomly interleaved with static gratings of the same SF and orientation. A few subjects enrolled during the initial phase of the study (2 MCI, 1 AD and 1 healthy subject) did not undergo this experiment. Further, we considered only those subjects for analysis in SSVEP experiment who had analyzable data for the Gamma experiment (see Artifact Rejection section below). This gave us a total of 221/12/5 subjects (99/2/2 females) for healthy/MCI/AD categories for the SSVEP experiment.

We presented each stimulus ∼30-40 times for both the Gamma and SSVEP experiments according to the subjects’ comfort and willingness. The entire recording session lasted for ∼30 minutes. Gamma experiment lasted for ∼25 minutes, with 1-2 short breaks (for 3-5 minutes) between blocks. This was followed by the SSVEP experiment (for ∼5 minutes) completed in one block. Unless otherwise stated, stimulus presentation of a particular orientation and spatial frequency is referred to as a “stimulus repeat” in this paper.

### Eye position analysis

We recorded eye signals (pupil position and diameter data) using EyeLink 1000 (SR Research Ltd) for all subjects (except for one subject each in healthy/MCI/AD categories). Eye-data for Gamma experiment is shown in Supplementary Figure 3. We rejected stimulus repeats with fixation breaks (eye-blinks or shifts in eye-position outside a square window of width 5° centered on the fixation spot) during −0.5s to 0.75s of stimulus onset (mean±SD: 16.8±14.2% and 19.1±15.2% for Gamma experiment; and 16.7±15.1% and 17.6±18.9% for SSVEP experiment, for healthy subjects and cases respectively). For the remaining repeats, all the subjects were able to maintain fixation with a standard deviation of less than 0.5° (healthy) and 0.6° (cases) for Gamma experiment and 0.6° (healthy subjects and cases) for SSVEP experiment in either directions.

### Artifact rejection

We used a pipeline to reject artifact-containing data as described in Murty et al. *(17)*. Briefly, we applied a repeat-wise thresholding process on both time-domain waveforms and multi-tapered PSD (between −500 ms to 750 ms of stimulus onset) to select bad repeats across electrodes. We discarded those electrodes that had more than 30% of all repeats marked as bad, and subsequently labelled any repeat as bad if it occurred in more than 10% of total number of remaining electrodes. We next discarded those electrodes that had PSD slopes (calculated in 56 Hz to 84 Hz range as described in *17*) less than 0. Finally, we discarded any block that did not have at least a single clean bipolar electrode pair (see Data Analysis subsection below) in any of the following three groups of bipolar electrodes: PO3-P1, PO3-P3, POz-PO3; PO4-P2, PO4-P4, POz-PO4 and Oz-POz, Oz-O1, Oz-O2. Despite these strict criteria, we ended up rejecting only 53/497 and 5/46 blocks for healthy subjects and cases; and we rejected only 5.5±6.4% and 4.9±3.2% of electrodes for healthy subjects and cases amongst those blocks that were analyzed. We then pooled data across all good blocks for each subject for final analysis. Those subjects who did not have any analyzable blocks (9/236 and 1/21 for healthy subjects and cases respectively) were discarded from further analysis. The total number of repeats that were finally analyzed were 276.2±87.2 for healthy subjects and 246.9±72.8 for cases.

A similar procedure was used for SSVEP experiment yielding 30.2±6.9 and 30.7±8.4 repeats for healthy subjects and cases respectively. Note that this experiment was always done towards the end, and therefore the signal quality could be poorer than the Gamma experiment. Consequently, we rejected 24/227 and 6/18 blocks for healthy subjects and cases; and 6.6±7.2% and 9.0±6.2% of electrodes amongst those blocks that were analyzed. Hence, we rejected data from 24/3/2 subjects out of 221/12/5 healthy/MCI/AD subjects as they did not have any analyzable blocks, leaving 197 healthy (93 female), 9 MCI (1 female) and 3 AD (1 female) participants for analysis for the SSVEP experiment.

### EEG data analysis

For all analyses we re-referenced data at each electrode offline to its neighboring electrodes (bipolar reference). We thus obtained 112 bipolar pairs out of 64 unipolar electrodes *(17)*. We considered the following bipolar electrodes for analysis: PO3-P1, PO3-P3, POz-PO3, PO4-P2, PO4-P4, POz-PO4, Oz-POz, Oz-O1, Oz-O2, which are inside the black encapsulation shown in Figure 1d. We discarded a bipolar electrode if either of its constituting unipolar electrodes was marked bad during artifact rejection. Data was pooled for the rest of the bipolar electrodes for further analysis.

We analyzed all data using custom codes written in MATLAB (The MathWorks, Inc, RRID:SCR_001622) as described in Murty et al. *(17)*. We computed PSD and the time-frequency power spectrograms using multi-taper method with a single taper using Chronux toolbox (*50*, http://chronux.org/, RRID:SCR_005547). We chose baseline period between −500 ms to 0 ms of stimulus onset, while stimulus period between 250 ms to 750 ms to avoid stimulus-onset related transients, yielding a frequency resolution of 2 Hz for the PSDs. We calculated time frequency power spectra using a moving window of size 250 ms and step size of 25 ms, giving a frequency resolution of 4 Hz.

We calculated change in power in alpha rhythm and the two gamma rhythms as follows:

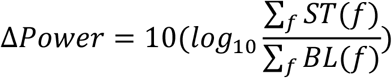

Where *ST* and *BL* are stimulus and baseline power spectra (across frequencies of interest, *f*) averaged across all analyzable repeats for all stimulus conditions and bipolar electrodes. For SSVEP experiment, we analyzed only the counter-phasing condition. Hence, we took the power at 32 Hz (twice the counter-phasing frequency, i.e. 16 cps) for analysis. Static gratings were presented in SSVEP experiment mainly to prevent adaptation and were discarded.

We generated scalp maps using the topoplot.m function of EEGLAB toolbox (*26*, RRID:SCR_007292), modified to show each electrode as a colored disc.

We calculated slopes (for Supplementary Figure 4) by fitting baseline PSD averaged across all analyzable repeats and bipolar electrodes with a power-law function using *fminsearch* in MATLAB: *P(f) = A. f*^−*β*^, where *P* is the PSD across frequencies *f, A* is scaling factor and *β* is the slope.

### Microsaccades and pupil data analysis

We detected microsaccades using a threshold-based method described earlier *(16)*, initially proposed by *(27)* for the analysis period of −0.5 s to 0.75 s of stimulus onset for the Gamma experiment. After removing the microsaccade-containing repeats, there were 128.1±71.1 (mean±SD, minimum 5) repeats for healthy subjects (n=226) and 115.4±43.2 (min: 30) for cases (n=18), excluding 3 participants for whom eye-data could not be collected. We used coefficient of variation (CV, ratio of standard deviation to mean) of pupil diameter across time for every repeat as a measure of pupillary reactivity to stimulus for that repeat *(17)*. We calculated CV for each analyzable trial separately and calculated mean CV across trials for every subject for comparison.

## Supplementary figures

**Supplementary Figure 1.**
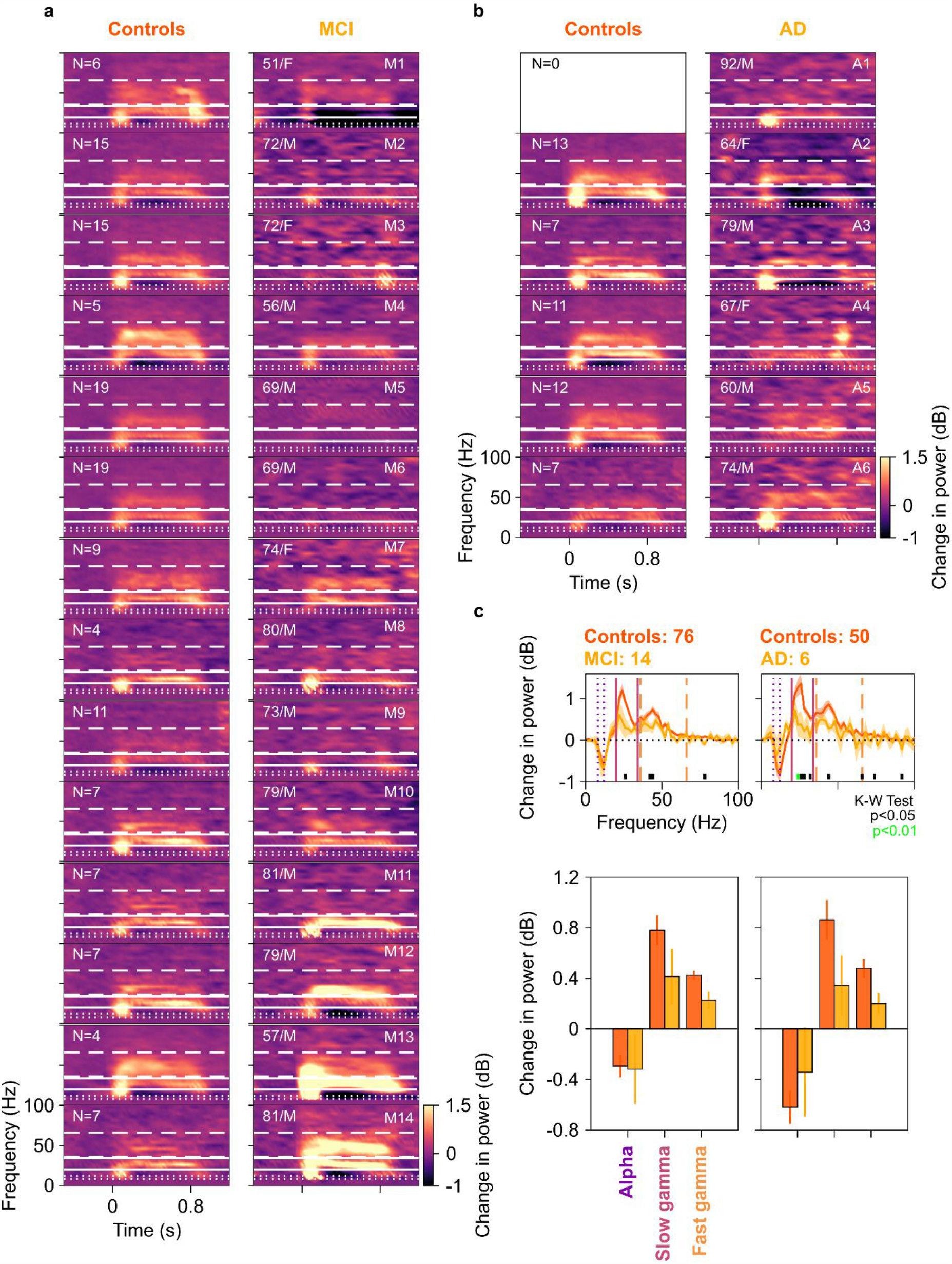
Alpha, slow gamma and fast gamma for MCI and AD compared to their corresponding controls. Median change in power spectrograms for individual MCI (a) and AD (b) cases plotted in right column and their healthy controls (left). The spectrograms are arranged by the difference in total gamma power (slow+fast, change from baseline in dB) for each MCI/AD case and the median total gamma power of their respective age and gender matched controls, highest difference plotted first. Age, gender, labels of cases and number of matched controls (N) are noted on spectrograms. White horizontal lines represent alpha (dotted), slow gamma (solid) and fast gamma (dashed) bands. Out of 14 MCI cases, only 3 (M11, M13-14) had higher slow gamma and 2 (M12, M14) had higher fast gamma than their controls. Similarly, out of 5 AD cases who had corresponding controls, only 1 (A6) had higher fast gamma than their controls. For the rest, MCI/AD cases had lower slow and fast gamma power than the median power for their healthy controls. c) Change in PSD (top row), and median change in power in alpha, slow gamma and fast gamma bands (bottom row), for MCI (left) and AD (right) cases (plotted in yellow) and their healthy controls (dark orange). Same format as Figures 1a and 1c. While the trends were similar to Figures 1a and 1c, they were not significant at a Bonferroni-corrected significance level of 0.017 (0.05/3; KW test, MCI vs controls: χ^2^(89)=2.06, p=0.15 for slow and χ^2^(89)=3.58, p=0.058 for fast gamma; AD vs controls, χ^2^(55)=5.19, p=0.02 for slow and χ^2^(55)=3.64, p=0.057 for fast gamma). This was probably because we used very stringent conditions for computation of gamma power, similar to our previous work *(16, 17)*. For example, for all participants, we used the same set of electrodes over which gamma was computed, as well as same time and frequency ranges. Further, we computed the total power within a band by simply summing the absolute power values within the band separately in baseline and stimulus periods and then taking a ratio. This estimate has larger contribution from lower frequencies in the band because of the power-law distribution of PSDs in baseline/stimulus periods. Consequently, if the traces are overlapping at lower frequencies within the band and diverge at higher frequencies, which was the case in the slow gamma range for both MCI and AD groups compared to their healthy controls, the total change in power in the band may not be significantly different. Therefore, our results could be improved by customizing the low frequency limit of the gamma band for each subject, as well as choosing only electrodes that show stronger gamma. For example, taking slow gamma range as 24-30 Hz improved p-values for both MCI (K-W test, χ^2^(89)=3.50, p=0.06) and AD (χ^2^(55)=6.74, p=0.009). We have refrained from such customization here because we wanted to study the efficacy of a simple and subject-independent computational procedure, but such data-driven subject specific optimization holds promise for improving the efficacy of a gamma-based biomarker. Moreover, there was a wide range of age of the MCI/AD cases for group-level analyses. As gamma power was shown to depend on age *(17)*, this could increase the variability in our data and hence adversely affect the observed significance. This problem was easily addressed by testing whether the median of differences in gamma power of individual cases with their controls is significantly different from zero, using paired Wilcoxon signed-rank tests in individual subject analysis (Figure 2). Finally, alpha power was not significantly different for MCI/AD compared to their controls (bar plots in panel 1c bottom row; KW test, MCI vs controls: χ^2^(89)=0.92, p=0.34; AD vs controls, χ^2^(55)=0.51, p=0.47).

**Supplementary Figure 2.**
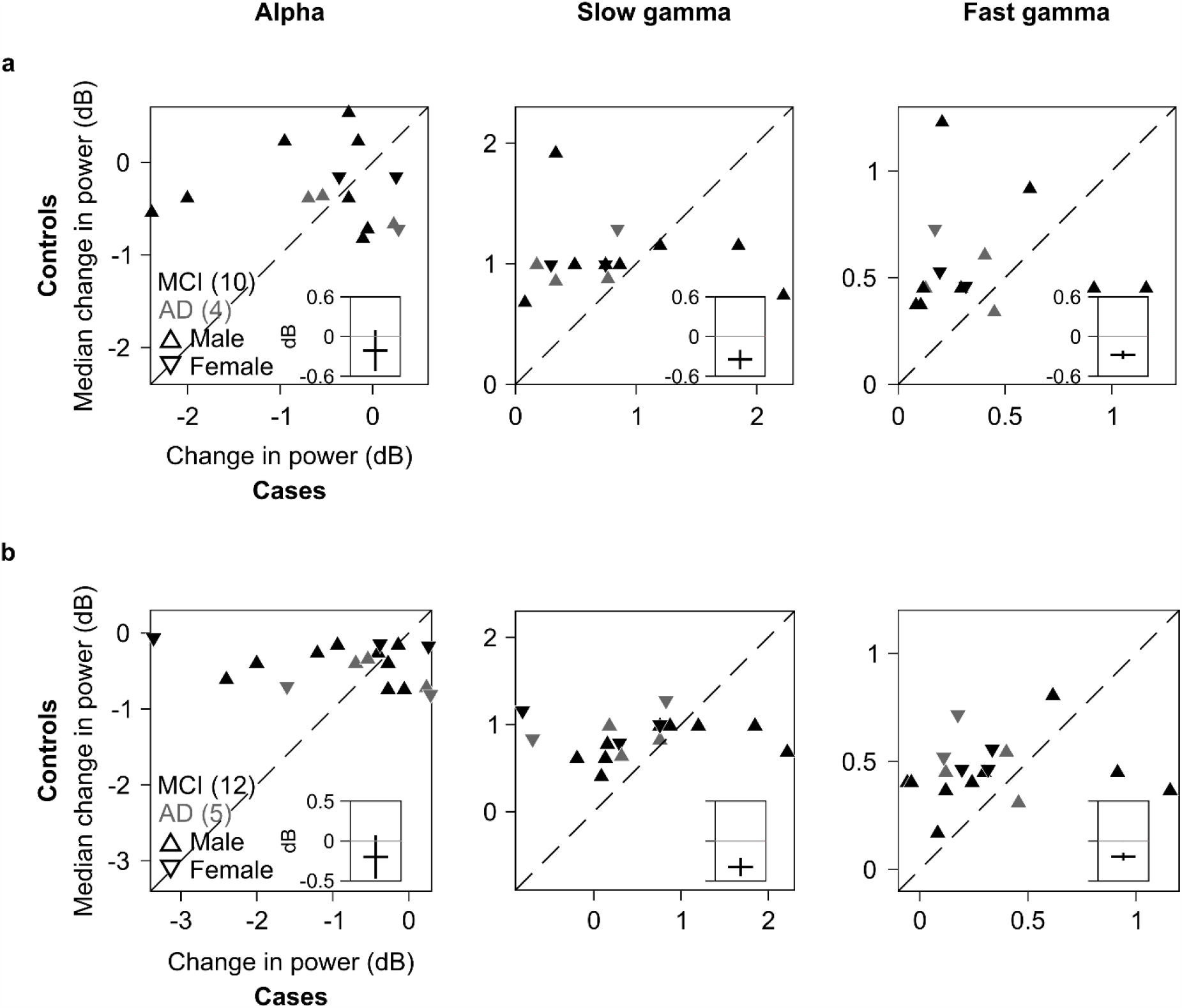
Additional comparisons between cases and corresponding healthy controls. Scatter plots showing change in power for cases (abscissa) and median change in power for corresponding healthy controls (ordinate) in alpha (right), slow gamma (middle) and fast gamma (left) bands. Same as in Figure 2. a) For those subjects with change in both slow and fast gamma power >0 dB (14 cases: 10 MCI and 4 AD and 96 controls). Left-tailed paired Wilcoxon signed-rank test: alpha, Z=-0.57, p=0.29; slow gamma, Z=-1.76, p=0.039; and fast gamma Z=-1.70, p=0.045. b) For those MCI/AD patients (N=17: 12 MCI and 5 AD; and their respective controls) whose diagnosis was confirmed by all 4 experts after the consensus diagnosis exercise (see SI Methods). Patients M4 and M8 were removed from analysis thus. Left-tailed paired Wilcoxon signed-rank test: alpha, Z=-1.09, p=0.14; slow gamma, Z=-2.08, p=0.019; and fast gamma Z=-1.99, p=0.023. Participant A1 (who also had change in both slow and fast gamma power >0 dB and whose diagnosis was confirmed during consensus diagnosis exercise) was excluded from both analyses above as he had no healthy control.

**Supplementary Figure 3.**
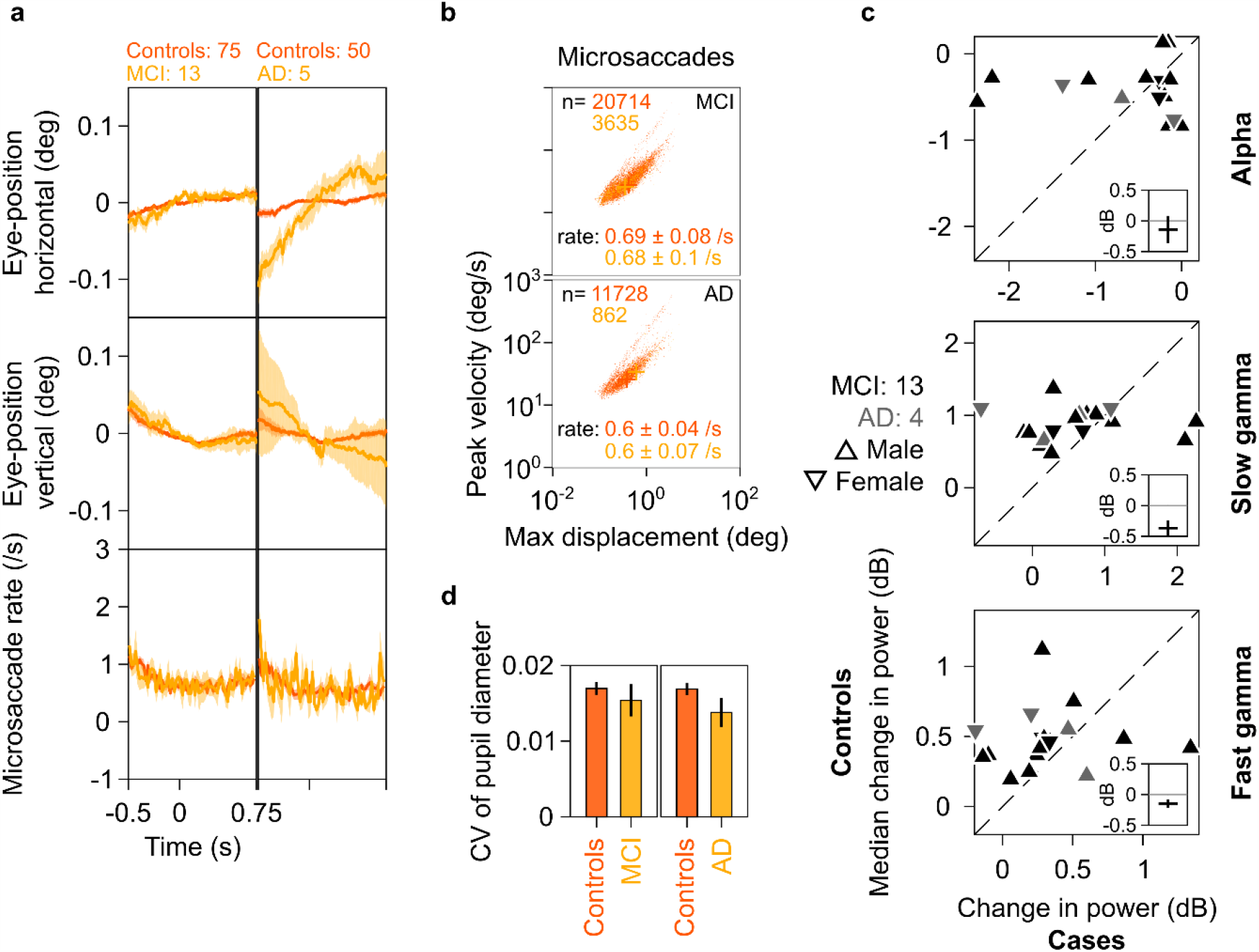
Eye position, microsaccades and pupillary reactivity for healthy/MCI/AD subjects. a) Left column: Eye-position in horizontal (top row) and vertical (middle row) directions; and histogram showing microsaccade rate (bottom row) vs time (−0.5-0.75 s of stimulus onset) for 13 MCI cases (yellow) and their 75 healthy controls (dark orange). Number of subjects in each age-group is indicated on top. Solid traces indicate medians, shaded patches represent ± SD of median after bootstrapping over 10,000 samples. Right column: same plots for 5 AD cases and 50 healthy controls. Eye position did not vary significantly across time between MCI/AD and control subjects except in the case of AD vs controls, where it varied slightly (but within ±0.1°). b) Main sequence plots showing peak velocity and maximum displacement of all microsaccades (number indicated on top) extracted from 13 MCI (top row), 5 AD (bottom row) cases indicated in yellow and their corresponding healthy controls (dark orange). Average microsaccade rate (median ± SD of median of 10,000 bootstrapped samples) across all subjects for each group is also indicated at the bottom of the panels. MCI/AD cases had similar microsaccade rates (also seen in panel 3a) and main sequence plots compared to their healthy controls. c) Scatter plots showing change in power for cases (abscissa) and median change in power for corresponding healthy controls (ordinate) in alpha (right), slow gamma (middle) and fast gamma (left) bands. Same format as in Figure 2, but stimulus repeats containing microsaccades have been discarded from analysis. Trends discussed in Figure 2 did not change qualitatively for these new set of repeats, as seen in these plots. Data of participant A1 has been discarded from analysis due to lack of healthy controls. d) Bar plots showing coefficient of variation of pupil diameter (reactivity of pupil to stimulus presentation, *see 17*) for 13 MCI (left), 5 AD (right) and their corresponding healthy controls. Height of bars indicate medians and error bars indicate ±SD of median of 10,000 bootstrapped samples. We did not find any significant difference between the MCI/AD and control groups in pupil reactivity as seen in the plot. These results ruled out potential biases due to ocular factors that could have influenced the results discussed in the text (Figures 1 and 2).

**Supplementary Figure 4.**
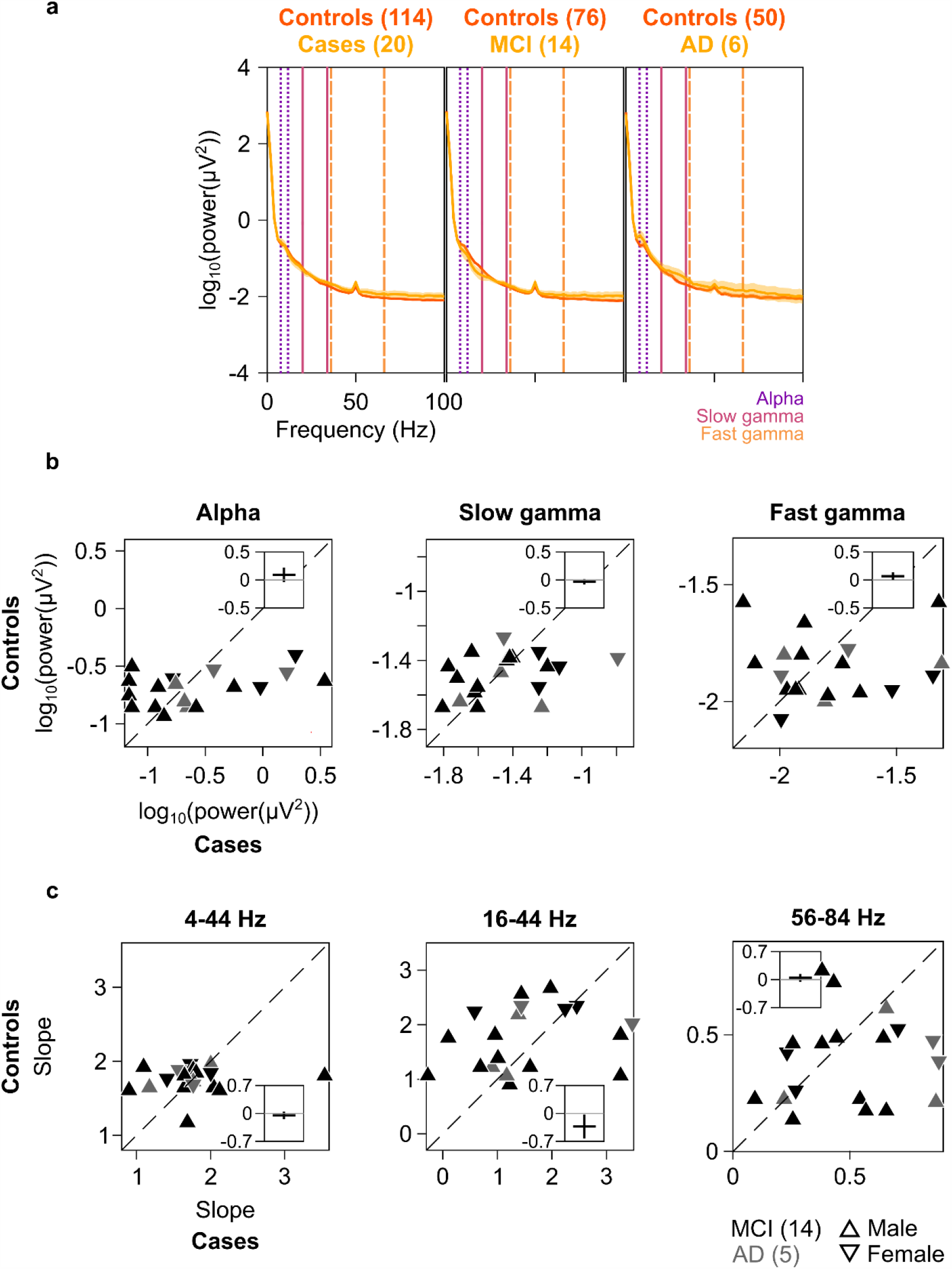
Baseline power and slopes in cases and healthy controls. a) PSDs for 20 cases (yellow) and 114 healthy controls (dark orange) are plotted in the left panel. Middle and right panels show these for MCI and AD groups separately (yellow), along with PSDs for corresponding healthy controls (dark orange). Same format as in Figure 1a. Median baseline PSDs for MCI/AD cases were overlapping with those of the control subjects. b) Scatter plots showing baseline absolute power (calculated in −500 – 0 ms of stimulus onset) for each of the 19 cases (abscissa) and median change in power for corresponding healthy controls (ordinate) in alpha (right), slow gamma (middle) and fast gamma (left) bands. c) Scatter plots showing baseline PSD slopes for each of the 19 cases (abscissa) and median change in power for corresponding healthy controls (ordinate). Frequency range considered for each scatter plot is mentioned above the plot. Panels (b) and (c) are in same format as Figure 2. Scatter was symmetrical across the identity line in both panels 4b and 4c (except for 16-44 Hz condition in panel 4c, where data points are slightly more above the identity line), suggesting that band-limited power in alpha, slow and fast gamma frequency ranges as well as slopes in 4-44 Hz and 56-84 Hz frequency ranges were comparable among cases and controls.

**Supplementary Figure 5.**
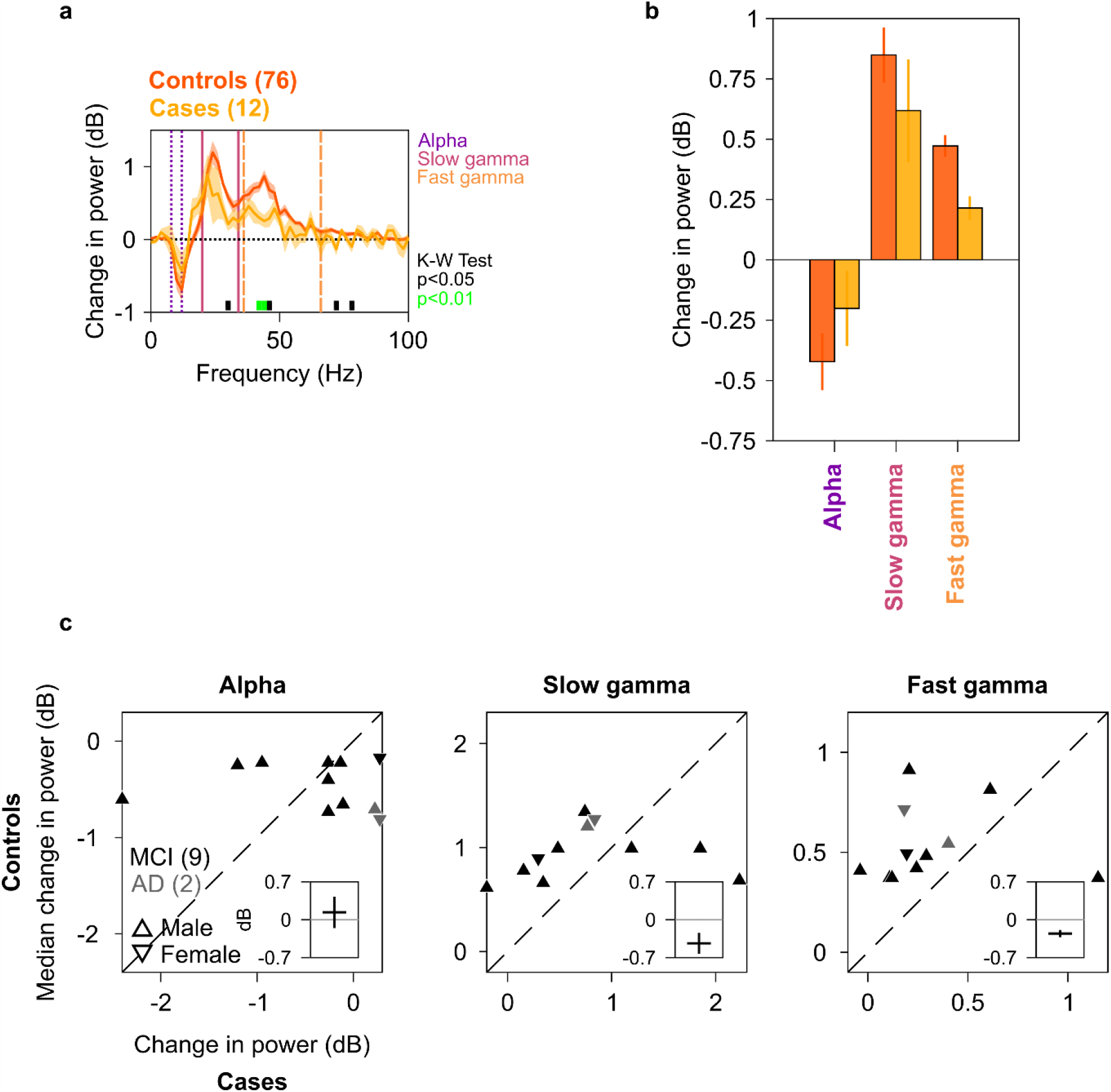
Alpha, slow and fast gamma in 12 cases and their healthy controls used for SSVEP analysis. Median change in PSD in (a), and median change in power in alpha, slow gamma and fast gamma bands in (b), for 12 cases (N = 9/3 for MCI/AD, plotted in yellow) and 76 healthy controls (dark orange; same set of cases and controls as in Figures 4a-d). Same format as Figures 1a and 1c. c) Scatter plots showing change in power for cases (abscissa) and median change in power for corresponding controls (ordinate) in alpha (right), slow gamma (middle) and fast gamma (left) bands. Same format as in Figure 2, but only for same set of subjects as in Figure 3e (N = 9/2 for MCI/AD).

**Supplementary Table 1:**
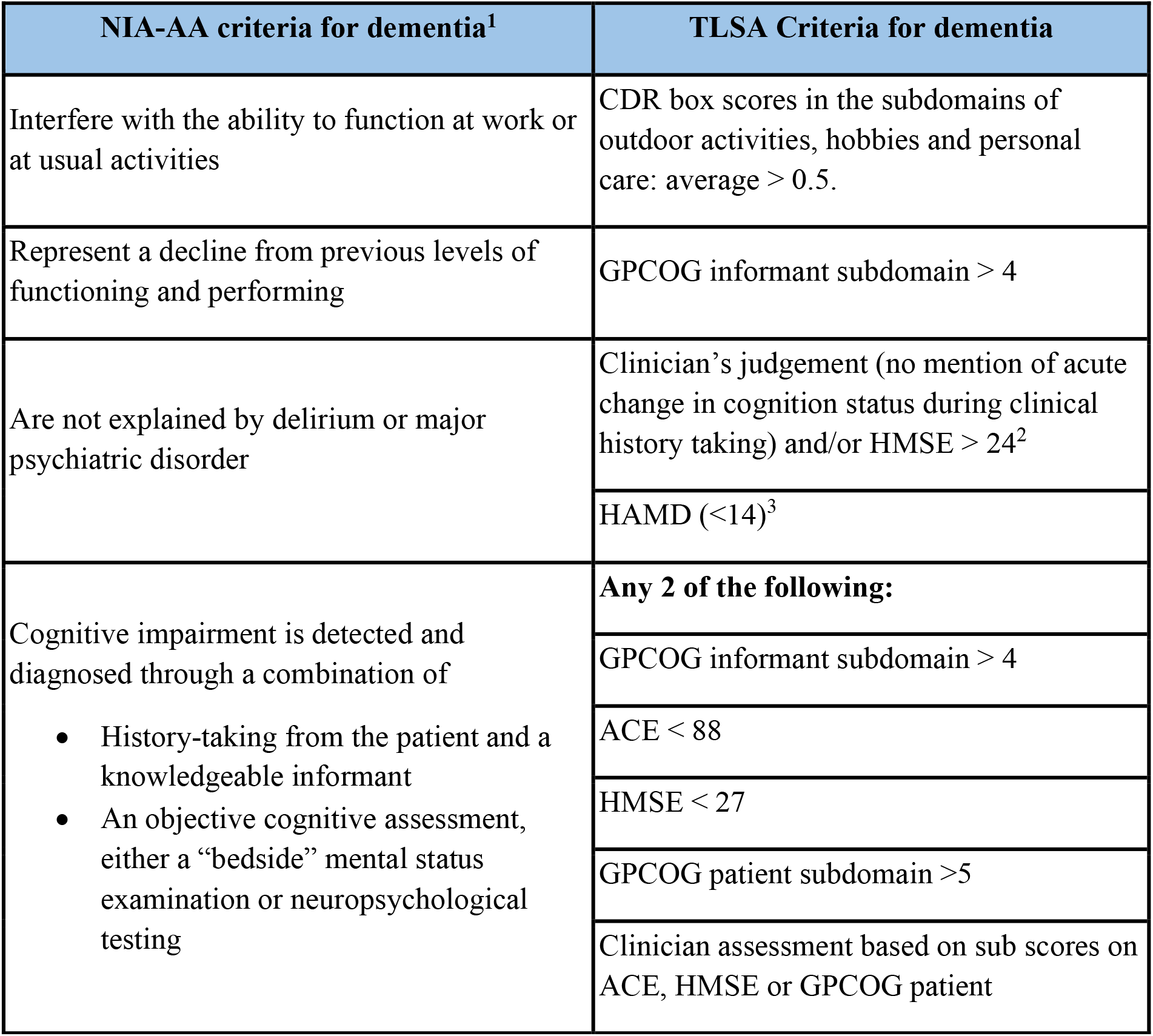
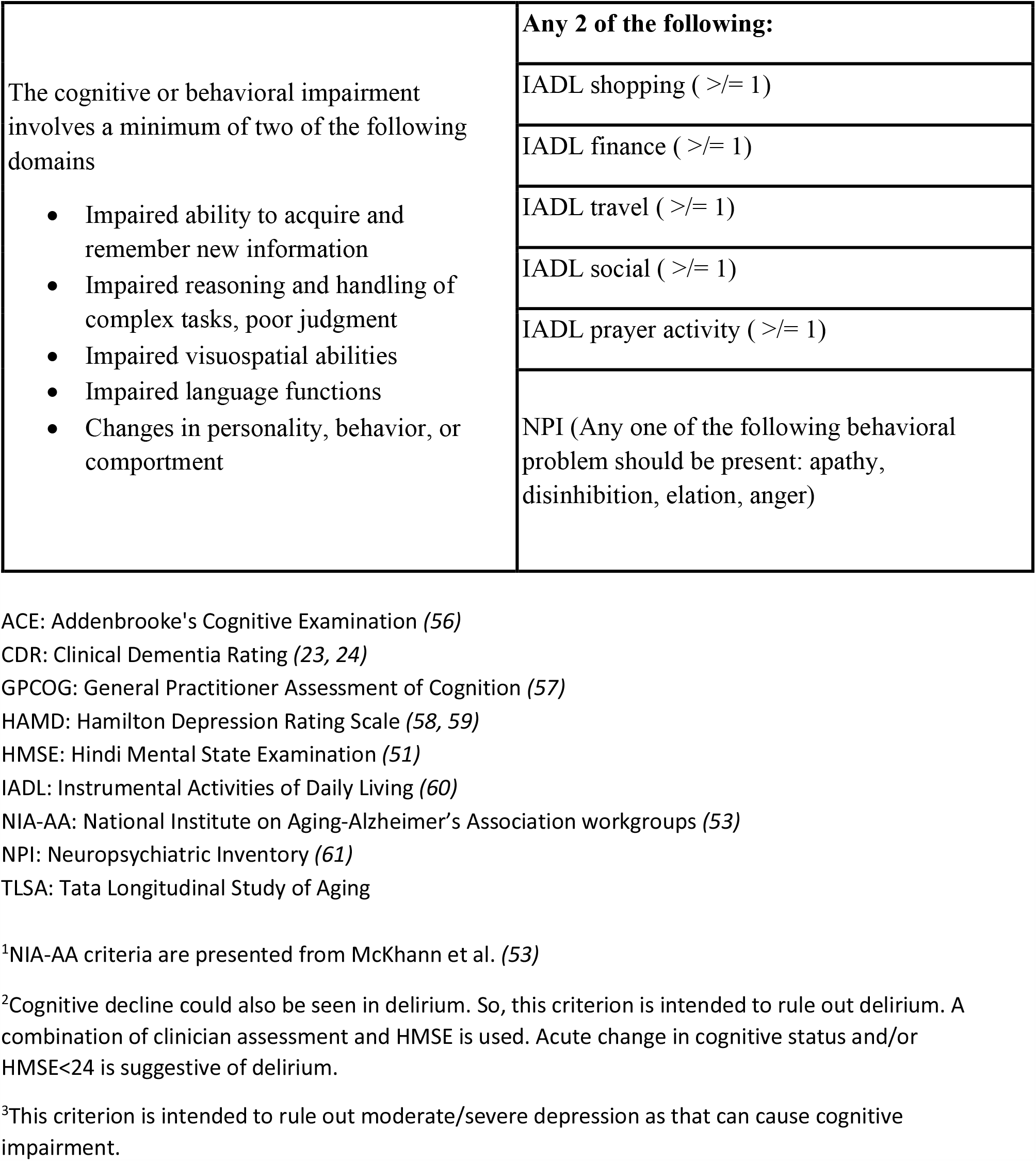
Criteria for dementia.

| NIA-AA criteria for dementia<sup>1</sup> | TLSA Criteria for dementia |
| --- | --- |
| Interfere with the ability to function at work or at usual activities | CDR box scores in the subdomains of outdoor activities, hobbies and personal care: average > 0.5. |
| Represent a decline from previous levels of functioning and performing | GPCOG informant subdomain > 4 |
| Are not explained by delirium or major psychiatric disorder | Clinician's judgement (no mention of acute change in cognition status during clinical history taking) and/or HMSE > 24<sup>2</sup> |
|  | HAMD (<14)<sup>3</sup> |
| Cognitive impairment is detected and diagnosed through a combination of<br>History-taking from the patient and a knowledgeable informant<br>An objective cognitive assessment, either a "bedside" mental status examination or neuropsychological testing | **Any 2 of the following:** |
|  | GPCOG informant subdomain > 4 |
|  | ACE < 88 |
|  | HMSE < 27 |
|  | GPCOG patient subdomain >5 |
|  | Clinician assessment based on sub scores on ACE, HMSE or GPCOG patient |
| The cognitive or behavioral impairment involves a minimum of two of the following domains<br>• Impaired ability to acquire and remember new information<br>• Impaired reasoning and handling of complex tasks, poor judgment<br>• Impaired visuospatial abilities<br>• Impaired language functions<br>• Changes in personality, behavior, or comportment | **Any 2 of the following:** |
| | IADL shopping ( $\geq 1$ ) |
| | IADL finance ( $\geq 1$ ) |
| | IADL travel ( $\geq 1$ ) |
| | IADL social ( $\geq 1$ ) |
| | IADL prayer activity ( $\geq 1$ ) |
|  | NPI (Any one of the following behavioral problem should be present: apathy, disinhibition, elation, anger) |

ACE: Addenbrooke's Cognitive Examination (56)
CDR: Clinical Dementia Rating (23, 24)
GPCOG: General Practitioner Assessment of Cognition (57)
HAMD: Hamilton Depression Rating Scale (58, 59)
HMSE: Hindi Mental State Examination (51)
IADL: Instrumental Activities of Daily Living (60)
NIA-AA: National Institute on Aging-Alzheimer's Association workgroups (53)
NPI: Neuropsychiatric Inventory (61)
TLISA: Tata Longitudinal Study of Aging

**Supplementary Table 2:**
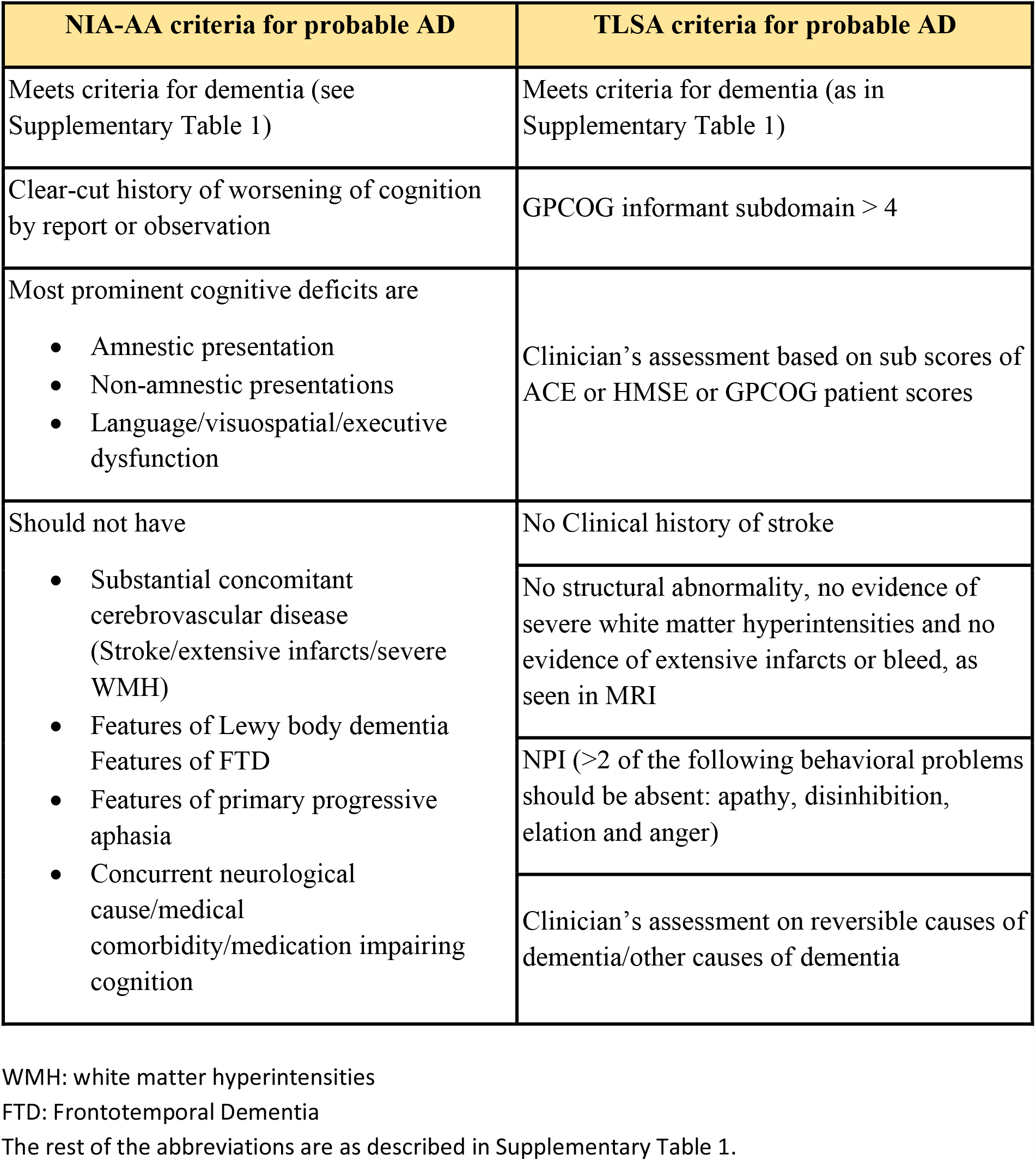
Criteria for probable AD.

| NIA-AA criteria for probable AD | TLSA criteria for probable AD |
| --- | --- |
| Meets criteria for dementia (see Supplementary Table 1) | Meets criteria for dementia (as in Supplementary Table 1) |
| Clear-cut history of worsening of cognition by report or observation | GPCOG informant subdomain > 4 |
| Most prominent cognitive deficits are<br>• Amnesic presentation<br>• Non-amnesic presentations<br>• Language/visuospatial/executive dysfunction | Clinician's assessment based on sub scores of ACE or HMSE or GPCOG patient scores |
| Should not have<br>• Substantial concomitant cerebrovascular disease (Stroke/extensive infarcts/severe WMH)<br>• Features of Lewy body dementia<br>• Features of FTD<br>• Features of primary progressive aphasia<br>• Concurrent neurological cause/medical comorbidity/medication impairing cognition | No Clinical history of stroke |
|  | No structural abnormality, no evidence of severe white matter hyperintensities and no evidence of extensive infarcts or bleed, as seen in MRI |
|  | NPI (>2 of the following behavioral problems should be absent: apathy, disinhibition, elation and anger) |
|  | Clinician's assessment on reversible causes of dementia/other causes of dementia |

WMH: white matter hyperintensities
FTD: Frontotemporal Dementia
The rest of the abbreviations are as described in Supplementary Table 1.

**Supplementary Table 3:** Criteria for MCI.

| NIA-AA criteria for MCI | TLSA criteria for MCI |
| --- | --- |
| Concern regarding a change in cognition | CDR = 0.5 |
| Impairment in one or more cognitive domains<br>• Memory<br>• Executive function<br>• Attention<br>• Language<br>• Visuospatial skills | **Any one of the following:** |
|  | GPCOG patient subdomain >5 |
|  | ACE < 88 |
|  | HMSE < 27 |
|  | Clinician's assessment based on sub scores of ACE/HMSE/GPCOG patient |
| Preservation of independence in functional abilities | IADL (average of finances, shopping, phone and meal preparation <0.5) |
| Not demented | CDR total score (<1) |
| Longitudinal decline in performance over repeated measures | 'Yes' for question no. 4<sup>1</sup> of CDR informant: memory subdomain **or** clinician assessment |

<sup>1</sup>Q4: Have there been some decline in memory in the past one year? (23, 24)
Abbreviations are as described in Supplementary Table 1.

